# Phenotyping extracellular vesicles and their serotonin transporter cargo in major depressive disorder

**DOI:** 10.1101/2025.02.05.25321729

**Authors:** Łukasz Zadka, Benjamin Eggerstorfer, Igor Buzalewicz, Chrysoula Vraka, Agnieszka Rusak, Godber M Godbersen, Agnieszka Opalińska, Jakob Unterholzner, Agnieszka Ulatowska-Jarża, Cecile Philippe, Katarzyna Haczkiewicz-Leśniak, Leo R Silberbauer, Matej Murgaš, Lukas Nics, Andreas Hahn, Marcus Hacker, Agnieszka Gomułkiewicz, Dan Rujescu, Marzenna Podhorska-Okołów, Rupert Lanzenberger, Gregor Gryglewski

## Abstract

With their ability to cross the blood-brain barrier, transport and regulate the release of various signaling molecules, extracellular vesicles (EVs) constitute a novel avenue to study intercellular communication in various pathologies, including psychiatric disorders. We studied 34 major depressive disorder (MDD) patients and 57 healthy controls and characterized EVs isolated from plasma using digital holographic tomography (DHT) and nanoparticle tracking analysis (NTA). EVs detected in DHT were markedly smaller and less variable in size in MDD patients. Furthermore, a lower variance of refractive indices related to the dry mass density of EVs was observed in patients. Meanwhile, NTA revealed a trend for elevated concentration of EVs in MDD. In DHT, EV dimensions correlated with plasma mRNA expression of exosomal markers *CD63* and *CD9*, with corresponding alterations in MDD. Levels of EV concentration and *CD9* were both negatively correlated with antidepressant treatment response. Utilizing electron microscopy and immunoblots we confirmed the presence of serotonin transporters (SERT) as EV cargo. SERT levels in EVs correlated with cerebral SERT binding potential in the amygdala measured using positron emission tomography. Next to the implication of altered EV communication in mood disorders, the pronounced differences in plasma EV characteristics and exosomal markers in MDD might inform the development of assays with diagnostic or prognostic value for clinical practice.

## Introduction

The complexity and heterogeneity of psychiatric disorders impede the development of biomarkers with diagnostic and prognostic utility (Abi-Dargham et al., 2023). Especially the clinical manifestation of depressive episodes displays a pronounced variety (Bailey et al., 2019). A possibility to complement our understanding of pathophysiological mechanisms of major depressive disorder (MDD) might be the study of extracellular vesicles (EVs), the role of which is only beginning to be characterized in the pathophysiology of psychiatric disorders (Saeedi et al., 2019).

EVs are phospholipid bilayer-delimited nano– and microparticles with a liquid content enriched in numerous bioactive molecules including proteins, nucleic acids, lipids, and labile molecules shielded from degradation in the extracellular environment. A conventional system divides EVs according to their dimensions into exomeres, exosomes, microbubbles, and oncosomes (Zadka et al., 2020a), whereas a simplified classification distinguishes small EVs (SEVs) for particles below 200 nm and large EVs (LEVs) for those over 200 nm in diameter (Théry et al., 2018). After endocytic incorporation of EVs, their cargo can change the biological activity and metabolism of distant target cells (Zhou et al., 2020). Importantly, EVs can traverse and integrate with the blood-brain barrier (Alvarez-Erviti et al., 2011; Ramirez et al., 2018). Given their capability to relay information from individual cells throughout the organism, EVs might play a pivotal role in intercellular communication in health and disease (Russo, 2017; Weihrauch et al., 2021; Zadka et al., 2020a). Their presence in bodily fluids, such as cerebrospinal fluid, blood, and urine, allows for non-or minimally invasive access to material for the isolation and analysis of EVs. This way of gaining tissue-specific information is being exploited in liquid biopsies, chiefly developed in cancer research, but holding particular promise for advancing precision psychiatry due to the limited access to brain tissue of patients.

A few recent studies characterized EVs isolated from blood samples of MDD patients and reported alterations in their cargo with regard to proteins (cytokines (Xie et al., 2023), mitochondrial proteins (Goetzl et al., 2021), brain derived neurotrophic factor (BDNF) (Gelle et al., 2021), insulin receptor substrate (Nasca et al., 2021)), mRNAs (Osborne et al., 2022), and micro RNAs (Honorato-Mauer et al., 2023; Jiang et al., 2021; Saeedi et al., 2021). While the selection of targets in some of these studies was informed by the inflammatory, neuroplasticity, and neurometabolic hypotheses of depression, so far the monoamine hypothesis, one of the mainstays of biological psychiatry, has received little attention in the EV field. This is despite the observations that alterations in mitochondrial proteins and reductions in size of EVs were reversed with response to treatment with selective serotonin reuptake inhibitors (SSRIs) (Saeedi et al., 2021). Indeed, further intriguing findings hint that studying EVs might help conflate the different theories by providing links between the systems involved: Serotonin was shown to stimulate the release of exosomes from microglia (Glebov et al., 2015), and, on the other hand, EVs released by gut bacteria altered the expression of proteins involved in the serotonergic system in the hippocampus (Yaghoubfar et al., 2020). While alterations in microRNA cargo of exosomes derived from individuals with MDD induced depressive-like behavior and decreased hippocampal neurogenesis in mice (Wei et al., 2020), exposure to BDNF was shown to alter the microRNA cargo of neuronal EVs such that they promoted excitatory synapse formation in recipient hippocampal neurons (Antoniou et al., 2023).

Here, we measured the size, concentration, and refractive indices of EVs isolated from patients with MDD using digital holographic tomography and nanoparticle tracking analysis. We related these parameters to plasma mRNA levels of exosome markers as well as to antidepressant treatment outcomes. Lastly, we established the presence of serotonin transporters (SERT) in EVs and correlated their levels with SERT binding measured using positron emission tomography in brain regions with known alterations in SERT in MDD (Gryglewski et al., 2014).

## Methods and materials

### Participants and study design

Patients diagnosed with MDD without antidepressant treatment in the three months prior to inclusion or comorbid psychiatric or somatic illness were recruited from the inpatient and outpatient clinics of the Department of Psychiatry and Psychotherapy at the Medical University of Vienna. Diagnosis was supported by the Structured Clinical Interview for DSM-IV for Axis I disorders (SCID-I). Data from 37 patients and 57 healthy controls was available for the reported analyses (Table 1). After inclusion, participants underwent PET/MR scans to measure SERT binding potentials (BP_P_) using [^11^C]DASB following established procedures detailed in the Supplementary Methods. After completion of scans, patients underwent a standardized open-label treatment algorithm starting with escitalopram 10 mg daily with biweekly follow-up visits for three months. Hamilton Depression Rating Scale (HAM-D), Beck Depression Inventory (BDI) and Montgomery-Åsberg Depression Rating Scale (MADRS) were used at screening and each follow-up visit. In case of non-response, defined by a reduction of at least 50% in the HAM-D 17, dosage was increased up to 20 mg escitalopram. In case of intolerable side-effects or non-response after 6 weeks, medication was switched to venlafaxine 75mg, mirtazapine 30mg or duloxetine 60mg. Further details on treatment can be found in previous publications (Klöbl et al., 2020; Seiger et al., 2021; Silberbauer et al., 2022). All participants provided written informed consent and received financial reimbursement for their participation. All procedures were reviewed and approved by the ethics committee of the Medical University of Vienna and carried out according to the Declaration of Helsinki. This study was registered before the start of recruitment at clinicaltrials.gov (NCT02711215).

**Table 1.** Demographics and clinical characteristics of study participants in the total sample and the subset standardized using International Society for Extracellular Vesicles (ISEV) criteria for further extracellular vesicle (EV) assessment (Witwer et al., 2013).

|  | Total sample set |  |  | Standardized subset |  |  |
| --- | --- | --- | --- | --- | --- | --- |
|  | MDD | HC | *p* | MDD | HC | *p* |
| Group size (n) | 34 | 57 |  | 20 | 20 |  |
| Age (years) | 28.3 ± 8.7 | 27.9 ± 8.3 | 0.82<sup>a</sup> | 26.1 ± 2.3 | 26.2 ± 3.5 | 0.87<sup>a</sup> |
| Sex (female/male) | 18/16 | 31/26 | 1<sup>b</sup> | 10/10 | 10/10 | 1<sup>b</sup> |
| Bodyweight (kg) | 68.2 ± 13.6 | 68.6 ± 11.5 | 0.89<sup>a</sup> | 68.7 ± 15.1 | 71.5 ± 14.5 | 0.54<sup>a</sup> |
| Body Mass Index | 22.8 ± 3.5 | 22.4 ± 2.5 | 0.59<sup>a</sup> | 22.9 ± 3.6 | 22.9 ± 2.8 | 0.87<sup>a</sup> |
| GPAQ total score | 14.8 ± 14.8<sup>#</sup> | 21.6 ± 24.5<sup>#</sup> | 0.14<sup>a</sup> | 14.4 ± 13.3<sup>#</sup> | 19.5 ± 19.0<sup>#</sup> | 0.40<sup>a</sup> |
| MADRS | 30.4 ± 5.9<sup>#</sup> | 0.5 ± 1.5<sup>#</sup> |  | 29.8 ± 4.3 | 1.1 ± 2.4<sup>#</sup> |  |
| HAM-D 17 | 22.5 ± 4.9 | 0.2 ± 0.6 |  | 21.7 ± 3.5 | 0.2 ± 0.5 |  |
| BDI | 27.6 ± 7.8<sup>#</sup> | 0.9 ± 1.7<sup>#</sup> |  | 27.6 ± 7.9 | 0.6 ± 1.0 |  |

MDD, major depressive disorder; HC, healthy control; GPAQ, Global Physical Activity Questionnaire; MADRS, Montgomery–Åsberg Depression Rating Scale; HAM-D 17, Hamilton Depression Rating Scale 17-item; BDI, Beck Depression Inventory. Continuous variables are shown as mean ± SD. <sup>a</sup> Independent two-sample *t*-test. <sup>b</sup> Chi-square test. <sup>#</sup> datapoint(s) missing.

### Availability of blood samples

After centrifugation at 3506 g for 4 minutes, plasma samples collected during PET/MR scans were stored at –80°C. All samples were transported on dry ice and next, gently thawed on ice prior the measurements. PAXgene Blood RNA tubes (catalog number: 762165, Qiagen) were used for collection of 2.5 ml venous blood during screening visits and stored at –20°C. Venous blood was not secured in 5 of 96 patients, and plasma was not collected in case of 3 patients. A total of 91 samples were available to study mRNA expression level of selected exosome markers, *CD9*, *CD63*, and *HSPA1A*, and SERT (*SLC6A4*).

To ensure the highest possible repeatability of the performed measurements, International Society for Extracellular Vesicles (ISEV) recommendations for sample normalization procedures were adapted (Witwer et al., 2013). Hence, we selected 20 patients and 20 healthy controls with equal sex ratios and the closest possible age range (20-33 years). The selected participants were Caucasian with a similar profile of physical activity that did not exceed 3 hours weekly and comparable BMI (Table 1).

### Isolation and precipitation of EVs

For procedures related to the isolation, precipitation, and imaging of EVs, the MISEV2018 criteria recommended by the ISEV were implemented (Théry et al., 2018). To confirm the exosomal nature of examined EVs additional staining against CD9 and CD63 antigens was used for transmission electron microscopy (TEM). An immunoblotting procedure was performed on a few randomly selected samples to detect the level of CD63 tetraspanin. Additionally, morphometric analyses of individual EVs using TEM imaging were performed. The total concentration of analyzed EVs and their size distributions were assessed using nanoparticle tracking analysis (NTA). Digital holographic tomography (DHT) was used to determine the refractive index (RI) and size distribution of EVs.

EV isolation was performed on 40 human plasma samples of 2000 µl volume each and on post-mortem frozen tissue of the human brain collected from the left and right occipital lobes during forensic autopsy as control (see Supplementary Methods). A total amount of 200 mg brain tissue was used for the precipitation of EVs and the isolation procedure was performed according to previously published protocols (Vella et al., 2017; Zadka et al., 2020b). After addition of a protease inhibitor cocktail, a differential centrifugation procedure was performed at 4°C as follows: 300 g for 5 minutes, 2,000 g for 10 minutes, and 10,000 g for 30 minutes. After each centrifugation cycle, the obtained supernatant was collected into new Eppendorf tubes. From the EV isolates obtained after centrifuging the samples at 10,000 × g, 750 µl of isolate was pipetted into new tubes and stored at –20°C until the analytic procedure was performed, and two Eppendorf tubes with a volume of 250 µl of isolate were used for precipitation of EVs. The ExoQuick procedure was used to prepare the exosomal precipitate by adding 63 µl of reagent (20 ml, catalogue number: 322-EXOQ20A-1, System Biosciences [SBI], California, United States) to each EV isolate obtained through centrifugation. The samples were then vortexed and incubated for one hour at 4°C. After incubation, a 30-minute differential centrifugation cycle was performed at 1500 g at 4°C. Then, the supernatant was removed and 100 µl of Dulbecco’s phosphate buffered saline (DPBS, 1000 ml, catalogue number: 14190136, Gibco, Thermo Fisher Scientific) was added to the obtained pellets. A total number of 120 samples (3 per participant) were prepared for NTA, DHT and immunoblotting measurements. Prepared pellets were used in the immunoblotting method to evaluate SERT isoforms present in the human brain and EV isolated from human brain and plasma.

### Nanoparticle tracking analysis (NTA)

NTA was used to detect and assess the trajectory of nanoparticles in EV isolates. The analysis was conducted at the Laboratory of Nanostructures, Institute of High Pressure Physics, Polish Academy of Sciences, Warsaw, Poland. The average size and size distribution of nanoparticles contained in the examined liquid fractions were analyzed. Measurements were performed using the NanoSight NS500 analyzer (serial number: 80336/A, Nanosight, Malvern Instruments Ltd, Software version 2.3.), and the light source was a diode laser with a wavelength of λ = 405 nm. NTA analysis was performed at room temperature (22.9 ± 0.3°C). All samples were diluted in PBS just before measurement. The instrument was calibrated with a standard solution in the form of Nanosphere Size Standard 200nm (catalogue number: 3200A). Measurements were made for nanoparticles in the size range of 10-1000 nm. The results constitute the mean values of the diameter of the tested EVs as well as their total concentration in units of 10^8^ nanoparticles/ml.

### Digital holographic tomography (DHT)

DHT is a new label-free technique that enables the assessment of cells visualized in 3D images based on the determined 3D-distribution of refractive index (RI) of the tested objects and measuring their physical and biological properties. The theoretical lateral and axial resolution of DHT systems is equal to 124 nm and 397 nm, respectively and provides morphological and biochemical information with sub-micrometer or even sub-nanometer resolution, which could make it suitable for examination of single EVs below the resolution limit of classical optical microscopy. In our recently published report, DHT was successfully used for the first time to detect and estimate the heterogeneity of individual EVs based on RI data (Zadka et al., 2021). For each EV precipitate, 20 3D-RI distributions consisting of 96 2D-RI tomograms were reconstructed by 3D Cell Explorer (Nanolive, Switzerland). From the obtained 3D-RI distributions, ranges of RI-values and the mean RI-value for each EV precipitate were determined. A probability density function based on the non-parametric distribution with normal kernel with support on (0, ∞) was fitted to each RI-histogram. The 2D-RI tomograms with the digitally stained EVs (based on their RI-values) were transformed into binary masks, and then EV rich regions of interests (area of their maximum cross-sections) were extracted using the Analyze Particles plugin in ImageJ software, followed by the determination of radius value of each examined EV. In total, nearly 400,000 EVs were examined, and their radii determined. Histograms of the distribution of the radius of the EVs were prepared, and a probability density function (pdf), based on inverse gaussian distribution (or Wald distribution) with the support on (0, ∞), was fitted to determine the average radius of the EVs for each participant. 2D-RI tomogram processing was performed in MATLAB R2020a (MathWorks, Inc., USA), except for the extraction of the EV regions, automatic counting, and determination of the EV size, which were performed using ImageJ software version 1.53i (NIH, Bethesda, MD; https://imagej.nih.gov/ij).

### Transmission electron microscopy (TEM)

TEM was performed to verify the exosomal origin of analyzed EVs. The purified exosomal pellets isolated through the ultracentrifugation procedure from human plasma samples were suspended in phosphate buffer saline (PBS; pH 7,4). Then, the buffer drop containing EVs was gently taken out from the ultracentrifuge tube and placed for 60 minutes on the shiny side of nickel grids, coated with formvar-carbon (200 mesh, Ted Pella, Redding, Ca, USA). All procedures were performed on the top of the droplet of each reagent’s solutions. Subsequently, the grids were washed 3 times by brief contact with PBS without magnesium and calcium ions (GenoPlast Biochemicals, Rokocin, Polska). Then, the EVs were fixed in 2% paraformaldehyde solution in PBS (Boster Biological Technology, Pleasanton, CA, USA) for 10 minutes and subsequently, the fixative was rinsed three times for 60 seconds. Next, double immunogold labelling was performed. In the first step, the incubation of the grids with the mixture of two primary antibodies from different species was conducted for 1 hour: polyclonal rabbit anti-SERT, extracellular (dilution 1:20, Cat. No. AMT-004, Alomone Labs, Jerusalem, Israel), and mouse monoclonal anti-hCD63 (dilution 1:20, IgG, clone 460305, Cat. No. MAB5048, R&D Systems, Inc. Minneapolis, MN, USA), antibodies were used, diluted in 1% bovine serum albumin, BSA in PBS (Albumin fraction V, Carl Roth, Mannheim, Germany). Then, the samples were rinsed with 1% BSA for 2 minutes per step (3X). Subsequently, the grids were incubated with the corresponding secondary antibodies, conjugated with different-sized colloidal gold nanoparticles. The first secondary antibody (code 810.011, goat-anti-rabbit IgG, H&L, conventional immuno gold reagents, 10-nm, AURION Immuno Gold Reagents & Accessories, Wageningen, The Netherlands), prepared in 0.1% BSA in PBS (dilution 1:20) was applied onto the grids for 1 h (dark chamber) and was dedicated for anti-SERT. To remove the unbound first secondary antibody, sections were washed with 0,1% BSA in PBS (3X, 2 minutes). Then the samples were incubated for 1 h with the second secondary antibody dedicated for anti-hCD63 (code 825.022, goat-anti-mouse IgG, H&L, conventional immuno gold reagents, 25-nm, AURION), prepared in 0.1% BSA in PBS (dilution 1:20). A 30 μl droplet of each antibody was used per each step of incubation. Then, the grids were rinsed with PBS (3X, 1 minute) and one time with deionized water. The incubations with the secondary antibodies were protected from light. Post-fixation of the exosome pellet was performed in 2.5% glutaraldehyde solution in distilled water for 10 minutes (Serva Electrophoresis, Heidelberg, Germany). Then, the fixative was removed by washing the grids with deionized water 5 times for 2 minutes, followed by counterstaining of samples with filtrated 2% uranyl acetate (Serva Electrophoresis) for 15 minutes. After staining, the grids were gently rinsed in large droplets of distilled water (3X). The last step was embedding the grids in 0.13% methyl cellulose (viscosity 25 cP, Merck KGaA, Darmstadt, Germany), diluted in distilled water for 10 minutes. After the samples have dried, the EVs were visualized in TEM (JEM 1011, Jeol, Tokyo, Japan), at an accelerating voltage of 80 kV. The TEM micrographs were performed using the iTEM1233 imaging platform equipped with the Morada Camera (Olympus, Munster, Germany) at magnification ranging from 50 to 250K.

### Western blot

Exosomal pellets isolated from human plasma, brain, and from human microvessel endothelial cells (HMEC-1, see Supplementary Methods) as control, were lysed in radioimmunoprecipitation assay (RIPA) buffer. Tissue samples were homogenized in Tissue Protein Extraction Reagent (T-PER) with EDTA, cocktail protease inhibitor (Heat Protease Inhibitor Coctail×100) and 0.5 mM phenylmethanesulfonyl fluoride (PMSF, Thermo Fisher Scientific, Wilmington, DE, USA). Bicinchoninic acid (Pierce BCA Protein Assay Kit) and the NanoDrop1000 (Thermo Fisher) were used to determine the total protein level. Denaturation of lysates was carried out in sample buffer [250 mM tris(hydroxymethyl)aminomethane (TRIS) pH 6.8, 40% glycerol, 20% (v/v) β-mercaptoethanol, 0.33 mg/ml bromophenol blue, 8% sodium dodecyl sulfate (SDS)] for 5 min at 95°C. Sodium dodecyl sulfate – polyacrylamide gel electrophoresis (SDS-PAGE) separation in 10% polyacrylamide gel in Mini Protean 3 apparatus (Bio-Rad, Hercules, CA, USA) with 20 μg of protein per lane was performed according to Laemmli method. Protein transfer was performed for 1 h at 140V in tris-glycine buffer with 20% methanol and 0.05% SDS, with polyvinylidene difluoride (PVDF) membrane 0.45 μm (Immobilon, Millipore, Bedford, MA, USA). The membranes were blocked in 5% milk in 0.05% tris-buffered saline with Tween 20 (TBST) and the incubation with primary rabbit antibody directed against SERT (1:200, in 5% milk in 0.05 % TBST, AMT-004, Alomone Labs, Jerusalem, Israel), β-tubulin (1:1000, in 0.1% bovine serum albumin (BSA) in 0.1% TBST, ab6046, Abcam, Cambridge, UK), and goat CD63 (1:1000, in 5% milk in 0.05% TBST, EXOAB, System Bioscience (SBI), Palo Alto, CA, USA) was carried out overnight at 4°C. Incubation with SERT blocking peptide (BLP-MT004, Alomone Lab) was performed according to the manufacturer’s recommendation. Incubation with secondary antibodies conjugated with horseradish peroxidase (HRP) was carried out for 1 h at room temperature with donkey anti-rabbit antibody (1:6000, in 5% milk in 0.05% TBST, Jackson ImmunoResearch, Suffolk, UK) or donkey anti-goat antibody (1:20 000, in 5% milk in 0.05% TBST, EXOAB). The level of SERT was determined by semi-quantitative densitometric analyses normalized on total protein measured with Ponseau S staining (Millipore) (Diller et al., 2021; Goldman et al., 2016). Chemiluminescence reactions were performed with Luminata Classico or Forte Immobilon Western HRP substrate (Thermo Fisher) and visualized with ImageLab software (Bio-Rad) and ChemiDoc MP System (Bio-Rad) with exposure time from 1 sec to 1 min. The experiments were run in triplicates.

### Statistical analysis

Analysis was conducted in R, version 4.3.2. The normal distribution was estimated each time prior to selection of the appropriate statistic test by visual inspection of histograms and Shapiro-Wilks test. To meet the assumption of normality, variables that did not exhibit a normal distribution were subjected to a log or arcsine hyperbolic transformation prior to statistical analysis. Parametric tests were used only if both tested groups had a normal distribution. The differences between the two studied groups were assessed using unpaired t-tests or Mann-Whitney U test. Correlations between variables were assessed using Spearman correlation coefficients. The level of statistical significance was set at α < 0.05 (two-sided).

## Results

### Characterization of extracellular vesicles in major depression

Transmission electron microscopy of EV isolates from plasma showed spherical nano– and micro-sized structures corresponding morphologically to exosomes (50 − 150 nm) and microvesicles (150 − 1000 nm) abundantly labelled against CD63 tetraspanin indicative of exosomes, which was confirmed using Western Blot (Figure 1A).

**Figure 1:**
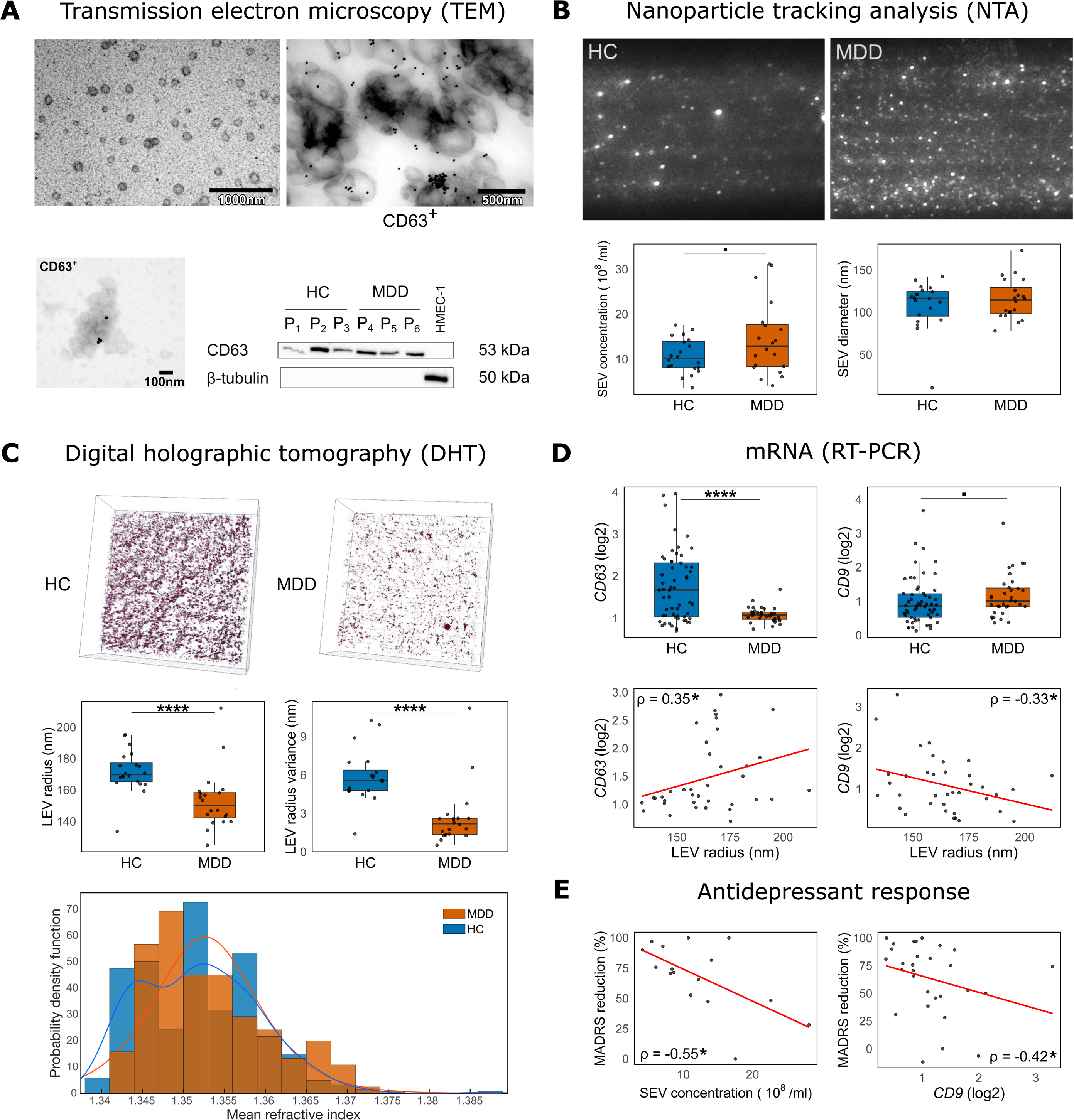
Characterization of extracellular vesicles isolated from plasma. **(A)** Transmission electron microscopy shows spherical nano– and micro-sized structures corresponding morphologically to exosomes (50 − 150 nm) and microvesicles (150 − 1000 nm) (upper left). Immunogold labelling particles against the exosomal marker CD63 tetraspanin supported the exosomal nature of examined EVs (right micrograph) in TEM micrographs of EVs isolated from a patient diagnosed with major depressive disorder (MDD). Western Blots confirmed the presence of CD63 in EV pellets from healthy controls (HC) and MDD patients. **(B)** Example images of CCD-camera used in Nanoparticle Tracking Analysis show differences in the number of nanoparticles detected in assessed cases. NTA revealed a trend toward higher small EV (SEV) concentration in MDD. **(C)** Representative rendered 3D-refractive index distributions of extracellular vesicles from one MDD patient and one HC captured using digital holographic tomography are shown. DHT was used to assess the RI values and the radius of large EVs (LEV). Lower EV radius and variance in size were observed in MDD. RI variance was higher in HC with a bimodal distribution. **(D)** Higher mRNA expression of *CD63* and lower expression of *CD9* were found in plasma of MDD patients. Expression of these exosomal markers was correlated with LEV radius. **(E)** Number of SEVs and *CD9* expression were correlated with antidepressant treatment response measured as the relative reduction in MADRS from screening to last follow-up visit. • p < 0.1, * p < 0.05, ** p < 0.01, **** p < 0.0001

In total, EV isolates from 40 participants (20 MDD patients) were included in NTA and DHT. NTA showed a trend towards higher total concentration of small EVs (SEVs) in the MDD group compared to healthy individuals (14.47 × 10^8^ ± 8.23 vs. 10.68 × 10^8^ ± 3.89 ml^-1^, t = –1.86, df = 27.09, p = 0.07, Figure 1B) with no significant differences in diameter of the particles detected between groups. Meanwhile, DHT which detects particles with a diameter of at least 187 nm revealed a lower mean radius of large EVs in patients compared to healthy controls (153.37 ± 19.16 vs. 171.75 ± 13.58 nm, W = 341, p < 0.0001, Figure 1C). Furthermore, the variance of EV radii captured by DHT was lower in MDD compared to controls (2.68 ± 2.39 vs. 5.89 ± 2.03 nm, W = 353, p < 0.0001). A similar constellation was observed with regard to the Refractive Indices of EVs: While no significant differences in average RI of EVs was observed between groups, the distribution of RI was bimodal in the control group and monomodal in MDD, with MDD lacking a putative subpopulation of EVs with lower RI, which was reflected by a lower variance in RI (1.87 × 10^-5^ ± 1.74 × 10^-5^ vs. 4.22 × 10^-5^ ± 2.66 × 10^-5^, t = 3.31, df = 32.72, p = 0.002). The RI values can be used to estimate the dry mass density (C) of EVs based on the formula:

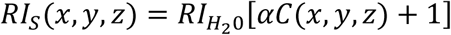

Considering that EV cargo may be composed of proteins, lipids, carbohydrates, and nuclei acid, we assumed an α constant equal to 0.0018 dL/g to estimate that the density of the low RI fraction (RI=1.34450) found in healthy controls is equal to 4.79 g/dL, while the average density of EVs from MDD patients was 8.482 g/dL (RI=1.35335).

### Markers for extracellular vesicles in MDD

To further corroborate our findings of alterations in EV in MDD, we measured mRNA expression of established EV markers (*CD63*, *CD9* and *HSPA1A*) in plasma of 91 participants (Figure 1D). There was a pronounced reduction in expression of *CD63* in MDD patients (1.08 ± 0.18 vs. 1.74 ± 0.82, t = 5.63, df = 75.37, p < 0.0001). We found a correlation between *CD63* expression and LEV radius (ρ = 0.35, p = 0.03) that was in line with the reduction of LEV radius in MDD. Furthermore, *CD9* expression was increased in plasma of MDD patients (1.10 ± 0.51 vs. 0.88 ± 0.50, t = –1.96, df = 84.64, p = 0.05) and was negatively correlated with LEV radius (ρ = –0.33, p = 0.04). Accordingly, *CD63* correlated negatively with *CD9* expression (ρ = –0.31, p = 0.002). We found no alterations in HSPA1A between groups or correlations between mRNA expression and other EV characteristics measured using NTA or DHT.

### Extracellular vesicles and clinical outcome

In an exploratory analysis we found that SEV concentration (ρ = –0.55, p = 0.03) and *CD9* expression (ρ = –0.42, p = 0.02) were negatively correlated with antidepressant treatment response measured as the relative reduction in MADRS from screening to last follow-up visit (Figure 1E).

### Serotonin transporters in exosomes and correlation with cerebral binding potentials

Immunoblots of EVs isolated from post-mortem brain showed discrete binding of anti-SERT antibody that was not present in reactions with SERT blocking peptide, indicating the presence of SERT as EV cargo in brain (Figure 2A). Following antibody validation, TEM was used to detect the presence of SERT binding in EV isolates positive for CD63 tetraspanin (Figure 2B). Western Blots performed in four cases showed that SERT protein was enriched in EV pellets isolated from plasma compared to total plasma (Figure 2C). Furthermore, a strong correlation of mRNA expression in plasma of *CD63* and *SLC6A4* coding for SERT was observed (ρ = 0.58, p < 0.0001) that was consistent with lower *SLC6A4* expression in MDD compared to healthy controls (4.09 ± 3.00 vs. 7.58 ± 6.04, t = 3.79, df = 69.51, p < 0.001; Figure 2D).

**Figure 2:**
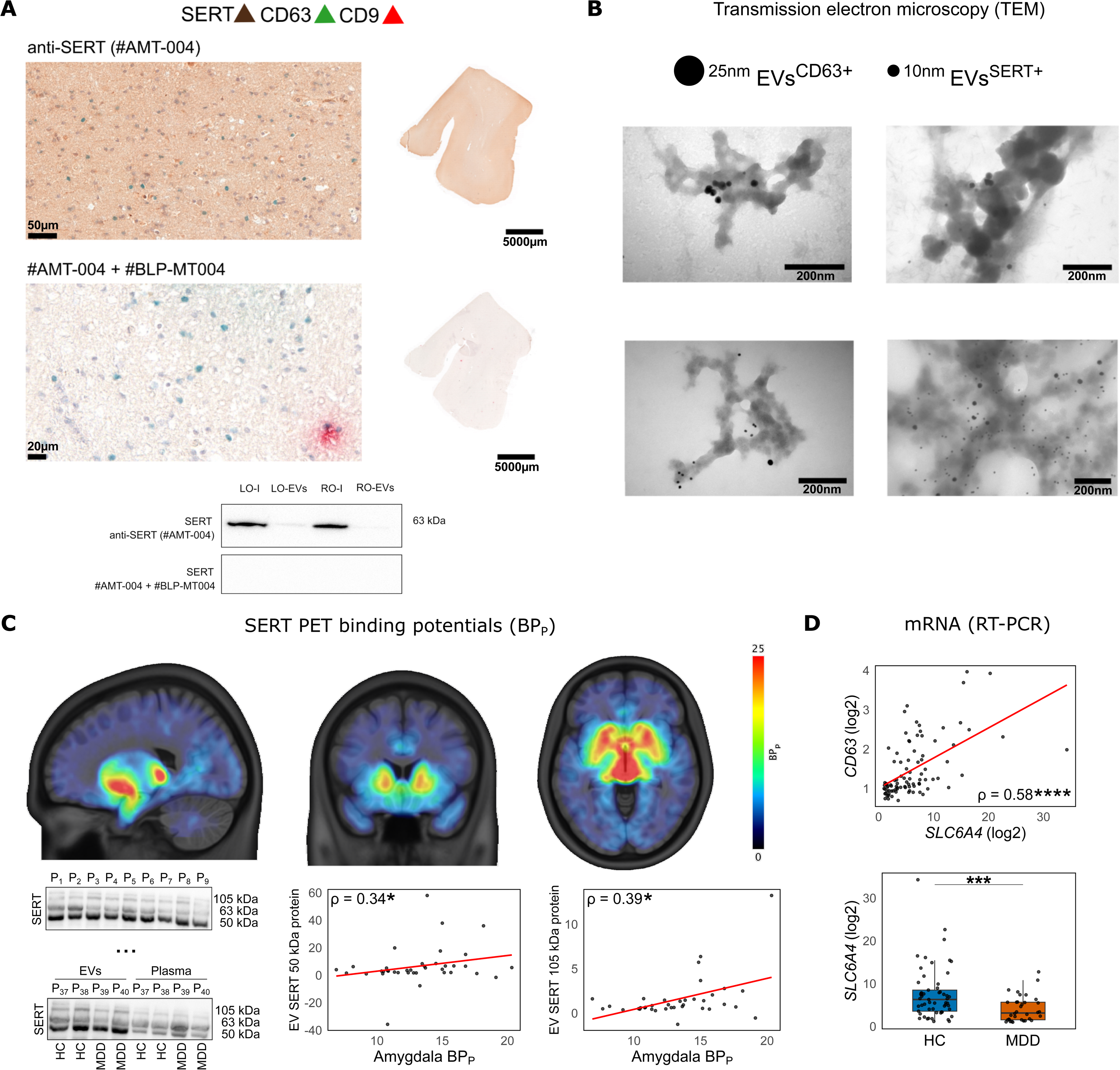
Analysis of serotonin transporter carriage of extracellular vesicles. **(A)** Post-mortem human brain sections of the occipital lobe were used as positive and negative controls for SERT immunostaining. Multicolor immunohistochemical reactions were performed against SERT, CD9 and CD63 antigens. SERT expression shown in brown (top) was abolished with the addition of blocking peptide (#BLP-MT004, bottom). Additionally, Western blot reactions were performed on tissue collected from the human left (LO) and right (RO) occipital lobe. For these samples, anti-SERT binding was independently confirmed on brain solid tissue homogenates (LO-I/RO-I) and EVs isolates from the same regions (LO-EVs/RO-EVs). **(B)** Transmission electron microscopy with double immunogold labelling showed expression of CD63 tetraspanin and SERT on EV isolates from plasma. **(C)** [^11^C]DASB binding potential (BP_P_) averaged for all participants is shown in the top row. Western blots showed SERT protein in EV pellets in three different lanes (50 kDa, 63 kDa, 105 kDa). Contrary to non-centrifuged plasma samples assessed in four cases, EVs pellets precipitated from the same plasma showed different protein level for SERT with higher SERT 50 kDa bands (bottom left). [^11^C]DASB uptake in the amygdala was correlated with EV SERT 50kDa and 105kDa protein expression. **(D)** *SLC6A4* mRNA expression in plasma correlates with *CD63* and is reduced in patients. * p < 0.05, *** p < 0.001, **** p < 0.0001

Having established the presence of SERT protein in EVs, we studied its relation to SERT binding in the brain measured using PET. Specifically, we correlated SERT BP_P_ of midbrain and amygdala, regions strongly implicated in MDD (Gryglewski et al., 2014; Schmaal et al., 2020), with SERT protein of different molecular weights (50, 63, 105 kDa) found in EVs (Figure 2C). Amygdala BP_P_ correlated with EV SERT protein 50 kDa (ρ = 0.34, p = 0.035) and 105 kDa lanes (ρ = 0.39, p = 0.017, which was robust to the exclusion of outliers (>2.5 SDs). No significant differences in EV SERT protein or SERT BP_P_ between MDD patients and controls were found.

## Discussion

We demonstrated altered EV morphology and quantity in MDD patients compared to healthy individuals. Patients had smaller LEVs with lower variance in size, whereas the mean diameters of SEVs were comparable. Nevertheless, a trend for higher SEV concentration in MDD was evident. This corroborates recent research showing reduced size and elevated concentrations of EVs in MDD (Saeedi et al., 2021). Moreover, LEV size and its variance displayed a noteworthy predictive value for MDD (Supplementary Figure S1). Obtained results suggest the presence of two subpopulations of low and high RI LEVs in controls and one population of high RI LEVs in MDD, implying reduced diversity of EVs in patients. These findings emphasize the proposed involvement of EVs in the pathophysiology of MDD and indicate functional changes in EV biogenesis that are common to both vesicle subclasses.

Although RT-PCR assessment of exosomal genes in plasma cannot replace the quantification of EVs, an overexpression of certain tetraspanins may suggest alterations in the release of EVs. CD9 and CD63 proteins are among most prominent targets in studies on EVs as they are involved in various cellular processes and in vesicle release from donor cells (Suárez et al., 2021). Accentuating the potential of *CD63* as a marker for EVs and suggesting exosomal involvement in MDD (Kalluri and LeBleu, 2020), *CD63* expression correlated positively with LEV size, with reductions noted in MDD compared to healthy controls. Furthermore, we found a trend for higher *CD9* levels in patients, along with a negative relationship with LEV size. CD63 and CD9 are also expressed in monocytes and macrophages in a mechanism depending on the activity of cytokines (Tippett et al., 2013), consistent with the inflammatory theory in MDD (Simon et al., 2021). Underpinning an exosomal influence in inflammation-driven depression, exosomes derived from depressive patients have been shown to reduce depressive-like behavior in animal models of depression (Wang et al., 2021), while inducing depressive-like behavior in normal mice (Wei et al., 2020). Nonetheless, pinpointing a well-defined origin of EV release responsible for fluctuations in mRNA expression proves challenging as other sources were tested positive for *CD9* and *CD63* (Bailey et al., 2011; Israels and McMillan-Ward, 2010).

Alterations in *SLC6A4* expression were associated with changes in central serotonergic transmission (Wang et al., 2012), and may directly influence cerebral SERT expression (Yaghoubfar et al., 2020). Our research revealed noteworthy reductions in *SLC6A4* mRNA levels in MDD and a strong positive correlation with the exosomal marker *CD63*. Additionally, we confirmed the presence of SERT protein in EV cargo through TEM and Western Blot analysis. Moreover, three distinct bands for SERT with different molecular weights were investigated: 50 kDa, 63 kDa and 105 kDa. In the brain, 50 kDa and 64 kDa SERT are predominantly detected (Mclane et al., 2011; Nyarko et al., 2018). In blood plasma, SERT has been demonstrated to be present with a mass of approximately 100 kDa in nucleated white blood cells (Barkan et al., 2004). SERT undergo multiple post-translational modifications including glycosylation and phosphorylation (Baudry et al., 2019), which may affect the final protein mass and result in subtle differences in molecular weight. However, we did not find the SERT 50 kDa in the normal post-mortem human brain samples used for immunoblotting negative and positive controls and we did not identify group differences in mean SERT protein levels. Nevertheless, contrasting distribution patterns of SERT between groups imply greater diversity within MDD, potentially indicative for distinct subgroups. Despite the uncertainty surrounding the origin of EVs carrying SERT, serotonergic neurons are implicated in exosome release and EVs can modulate SERT expression (Glebov et al., 2015; Yaghoubfar et al., 2020), leading to the speculation that alterations of EV SERT in some patients is a result of a disbalance in serotonin homeostasis.

[^11^C]DASB PET is considered as the gold standard for SERT quantification in the human brain, and cerebral uptake proved to be diminished in several brain regions in MDD patients by miscellaneous studies (Gryglewski et al., 2017, 2014). We identified a relationship between [^11^C]DASB BP_P_ and SERT protein levels in peripheral EVs. The 50kDa and 105kDa SERT bands moderately correlated with radiotracer uptake in the amygdala, which may indicate a link between peripheral and cerebral SERT concentrations.

The prediction of outcome through biomarkers poses an important obstacle in the treatment of MDD. A study found that changes in miRNA cargo of neuron-derived EVs were associated with the efficacy of antidepressant treatment (Saeedi et al., 2021). Adding to these findings, the peripheral concentration of SEVs and *CD9* levels were related to the clinical outcome in the treatment of MDD with commonly prescribed antidepressants, measured by MADRS. However, *CD63* expression and other EV features were not linked to treatment outcome (Supplementary Figure S2).

Due to technical constraints inherent in the isolation of EVs, obtaining completely pure EV fractions is presently unfeasible. Regardless of the employed isolation method, the potential for sample contamination by vesicles of varying diameter ranges and diverse biogenesis pathways cannot be definitively eliminated (Zhang et al., 2019). NTA is one of the methods recommended by ISEV for the evaluation of EVs (Théry et al., 2018), however, with this technique all vesicles in a given size range are counted, which makes it impossible to exclude other particles with similar morphometric parameters (Takov et al., 2019). Notably, we did not enrich EV isolates for neuronal origin (Gomes and Witwer, 2022). However, commonly used markers for neuron-derived EVs are not entirely specific (Norman et al., 2021). Moreover, while NTA and DHT are restricted in their availability requiring specialized personnel and expensive equipment, analysis of mRNA expression of exosomal markers offers a readily available option in EV assessment.

In conclusion, we identified substantial changes in EV morphology and distribution of MDD patients compared to healthy individuals, uncovered a connection between EVs and serotonergic transmission in MDD and identified distinct relationships between exosomes and treatment response. Consequently, plasma EVs may represent an influential factor in the molecular neurobiology of MDD and further investigation on EVs in this context seems reasonable.

## ClinicalTrials.gov

Patient Stratification and Treatment Response Prediction in Neuropharmacotherapy Using Hybrid Positron Emmission Tomography/Magnetic Resconance Imaging (PET/MR); https://clinicaltrials.gov/study/NCT02711215; NCT02711215.

## Data Availability

All data produced in the present study are available upon reasonable request to the authors

## Abbreviations

EVs: extracellular vesicles
MDD: major depressive disorder
SERT: serotonin transporter
DHT: digital holographic tomography
NTA: nanoparticle tracking analysis
TEM: transmission electron microscopy
RT-PCR: Real-time polymerase chain reaction
BDI: Beck Depression Inventory
MADRS: Montgomery–Åsberg Depression Rating Scale
HAM-D: Hamilton Depression Rating Scale
RI: refractive index
GPAQ: Global physical activity questionnaire

## Acknowledgements

We would like to express our thanks to the patients who participated in this study. We thank Marius Hienert, Thomas Vanicek, Alexander Kautzky, Johannes Jungwirth, Paul Michenthaler, Alim Emre Başaran, Radheshyam Stepponat and the medical students of the Neuroimaging Labs for their medical support, Edda Winkler-Pjrek, Siegfried Kasper, Markus Hartenbach for medical supervision, and Georg Kranz for his assistance in randomization of participants. We are grateful to Karoline Einenkel, Elisa Sittenberger and Vera Ritter for their administrative support, and Gregory M. James and Murray B. Reed for technical support. We gratefully acknowledge the support of Verena Pichler, Neydher Berroterán-Infante, Theresa Balber, Markus Mitterhauser, Wolfgang Wadsak and Eva-Maria Klebermass in tracer synthesis and metabolite analysis and supervision.

## Author contributions

Conceptualization: ŁZ, GG, RL; Methodology: ŁZ, IB, AR, RL, GG; Software: ŁZ, IB, AO, MM, AG; Validation: ŁZ, BE, MPO, GG; Formal Analysis: ŁZ, BE, IB, AR, AO, AUJ, KHL, MM, AG, GG; Investigation: GG, GMG, JU, LRS, BE; Resources: ŁZ, IB, CV, CP, LN, MPO, RL, DR, MH, AH, LN; Data Curation: ŁZ, BE, IB, MPO, GG; Writing original-draft: ŁZ, BE, IB, AR, AUJ, KHL, AG, GG, RL; Writing review and editing: all authors, Visualization: ŁZ, BE, MM; Supervision: AH, MH, DR, GG, RL, MH, LN, AH; Project administration: RL, ŁZ, GG; Funding acquisition: RL, ŁZ.

