## Supplemental Materials for "Phenotyping extracellular vesicles and their serotonin transporter cargo in major depressive disorder"

### Supplementary materials

#### Supplementary methods

##### Imaging procedures and quantification of serotonin transporter (SERT) binding potentials

PET/MR scanning and quantification of SERT binding potentials ($BP_P$) were performed following an established protocol (Gryglewski et al., 2017). In a randomized cross-over design participants underwent two PET/MR scans with an interscan interval of at least three days. Citalopram (8 mg) or placebo (saline) was infused during either the first or the second scan. For the analyses reported in this work, imaging data and plasma samples collected during the placebo scan were used. On PET/MR scan days, arterial and venous cannulation were performed and application of the serotonin transporter specific tracer [$^{11}$C]-N,N-dimethyl-2-(2-amino-4-cyanophenylthio)-benzylamine ([$^{11}$C]DASB) was performed as bolus plus constant infusion. Citalopram 8mg or saline were infused over 8 minutes starting 70 minutes after initiation of tracer application. Arterial samples were collected into S-Monovettes 9 ml, Lithium-Heparin, 92x16 mm (catalog number: 02.1065, Sarstedt) at 2, 5, 8, 12, 18, 30, 50, 60 and 70 min after initiation of drug application.

PET data was acquired in listmode on a PET/MR scanner (Biograph mMR, Siemens Healthineers, Erlangen, Germany) and reconstructed using an ordinary Poisson-ordered subset expectation maximization algorithm (OP-OSEM, 3 iterations, 21 subsets). Attenuation correction was performed using a low-dose CT scan (Siemens Biograph TruePoint PET/CT) recorded on a separate occasion. PET data was registered to a T1-weighted image (MPRAGE, TE/TR=4.21/2000 ms, 1 × 1 mm in-plane resolution, 1 mm slice thickness, 0.1 mm gap) and normalized to Montreal Neurological Institute (MNI) space using SPM 12 (Wellcome Trust Center for Neuroimaging, London, United Kingdom). Activity of regions of interest defined by the Automated Anatomical Labeling atlas at equilibrium measured between 55 and 95 min after drug challenge was averaged and divided by the average metabolite-corrected plasma activity in that timeframe to obtain distribution volumes ($V_T$). Binding potentials ($BP_P$) were calculated by subtracting the distribution volume of cerebellar grey matter, which served as the reference region due to its low SERT binding (Tzourio-Mazoyer et al., 2002).

##### Human brain tissue

To validate antibodies used and confirm the presence of SERT in EVs isolated from human brain, tissue was obtained for immunohistochemical and immunoblotting reactions For this purpose, archived material was used, the collection of which was approved by the Bioethical Committee at Wroclaw Medical University (protocol number 665/2017).

The brain tissue material was collected in 2018 at the Department of Forensic Medicine in Wrocław, Poland, during forensic medical autopsy. A thorough macro- and microscopic evaluation was performed. The macroscopic hallmarks of the brain were carefully estimated. The microscopic examination was performed on the standardized histological slides using hematoxylin and eosin staining. No signs of significant tissue autolysis were found. Brain tissue was collected symmetrically, from both hemispheres and lobes. Two tissue fragments were obtained from the left and right occipital lobes (Brodmann area 17) of one donor (sex: male, age: 54, height: 176 cm, body weight: 112 kg, BMI: 36.2, cause of death: cardiac arrest (natural)). Fresh tissue sections of 0.5–1 cm in size were collected from each anatomical area and were kept frozen until further analysis. Before the collection to fresh tubes, the specimens were divided into two different tissue fragments and placed in individual embedding cassettes and then embedded in paraffin. Next, the Formalin-Fixed Paraffin-Embedded (FFPE) blocks were sectioned for immunohistochemistry.

*Triple stain multicolor enzymatic immunohistochemistry assay*

Whole histological slides 4 μm thick were sectioned from one FFPE block representing the right occipital lobe. To detect co-expression between SERT and exosomal markers three different primary antibodies were used as follows: rabbit polyclonal antibody against 4$^{th}$ extracellular loop of SERT (1:200, catalogue number: AMT-004, alomone), mouse anti-human anti-CD63 monoclonal antibody (1:100, catalogue number: MAB5048, R&D Systems) and mouse anti-human anti-CD9 monoclonal antibody (1:100, catalogue number: MAB25292-100, R&D Systems). To visualize the immunohistochemical staining the TripleStain IHC Kit: M&M&R on Human tissue (DAB, AP/Red & HRP/Green) was used (catalogue number: ab183286, abcam). All reactions were performed in accordance with the manufacturer protocol. For the negative control against SERT, an additional slide from the same FFPE block was preincubated with SERT (extracellular) blocking peptide (catalogue number: BLP-MT004, alomone).

**Cell culture**

Human microvessel endothelial cells line HMEC-1 were used as negative control for western blot analysis. HMEC-1 cells were obtained from ATCC (American Type Culture Collection ATCC, Old Town Manassas, VA, USA) and cultured in MCDB131 medium supplemented with epidermal growth factor (EGF) and hydrocortisone, L-glutamine (Thermo Fisher, Wilmington, DE, USA), penicyline-streptomycine solution and 10% FBS (Sigma-Aldrich St. Louis, MO, USA). Cell cultures were kept at 37°C, 5% CO2 and 95% humidity, the media were changed twice a week and cells were passaged when confluency reached about 70 %. Trypsin – EDTA ethylenediaminetetraacetic (EDTA) solution (Sigma-Aldrich) was used for cell trypsinization.

**Real-time polymerase chain reaction (RT-PCR)**

PAXgene Blood RNA tubes (catalog number: 762165, Qiagen) were used for collection of 2.5 ml venous blood during screening visits and stored at -20°C. Total RNA was extracted from studied blood samples with the PAXGene Blood RNA Kit (PreAnalytiX, Qiagen, Hilden, Germany) according to the manufacturer's protocol. The yield and purity of extracted RNA were measured using the NanoDrop1000 spectrophotometer (Thermo Fisher Scientific, Wilmington, DE, USA). The RNA integrity was analyzed by capillary electrophoresis using the Agilent 2100 bioanalyzer and the associated RNA 6000 Nano LabChip kit (Agilent Technologies, Palo Alto, CA, USA). Complementary cDNA was synthesized using the High Capacity cDNA Reverse Transcription Kit (Applied Biosystems, Carlsbad, CA, USA) as described in the protocol. The mRNA expression of *SLC6A4*, *CD63*, *CD9* and *HSPA1A* was determined by quantitative real-time PCR with 7900HT Fast Real-Time PCR System and TaqMan Gene Expression Master Mix (Applied Biosystems, Carlsbad, CA, USA). Glyceraldehyde 3-phosphate dehydrogenase (*GAPDH*) was used as the reference gene. The following sets of primers and TaqMan probes were used for the reactions: Hs00984356_m1 for *SLC6A4* (*SERT*), Hs01041238_g1 for *CD63*, Hs01124022_m1 for *CD9*, Hs00359163_s1 for *HSPA1A* (*Hsp70*) and Hs99999905_m1 for *GAPDH* (Applied Biosystems, Carlsbad, CA, USA). All measurements were performed in triplicates under following conditions: activation of polymerase at 50ºC for 2 minutes, initial denaturation at 94ºC for 10 minutes, followed by 40 cycles of denaturation at 94ºC for 15 s and annealing with elongation at 60ºC for 1 minute. The relative mRNA expression of studied markers was calculated with the ΔΔCt method.

#### Supplementary Figures

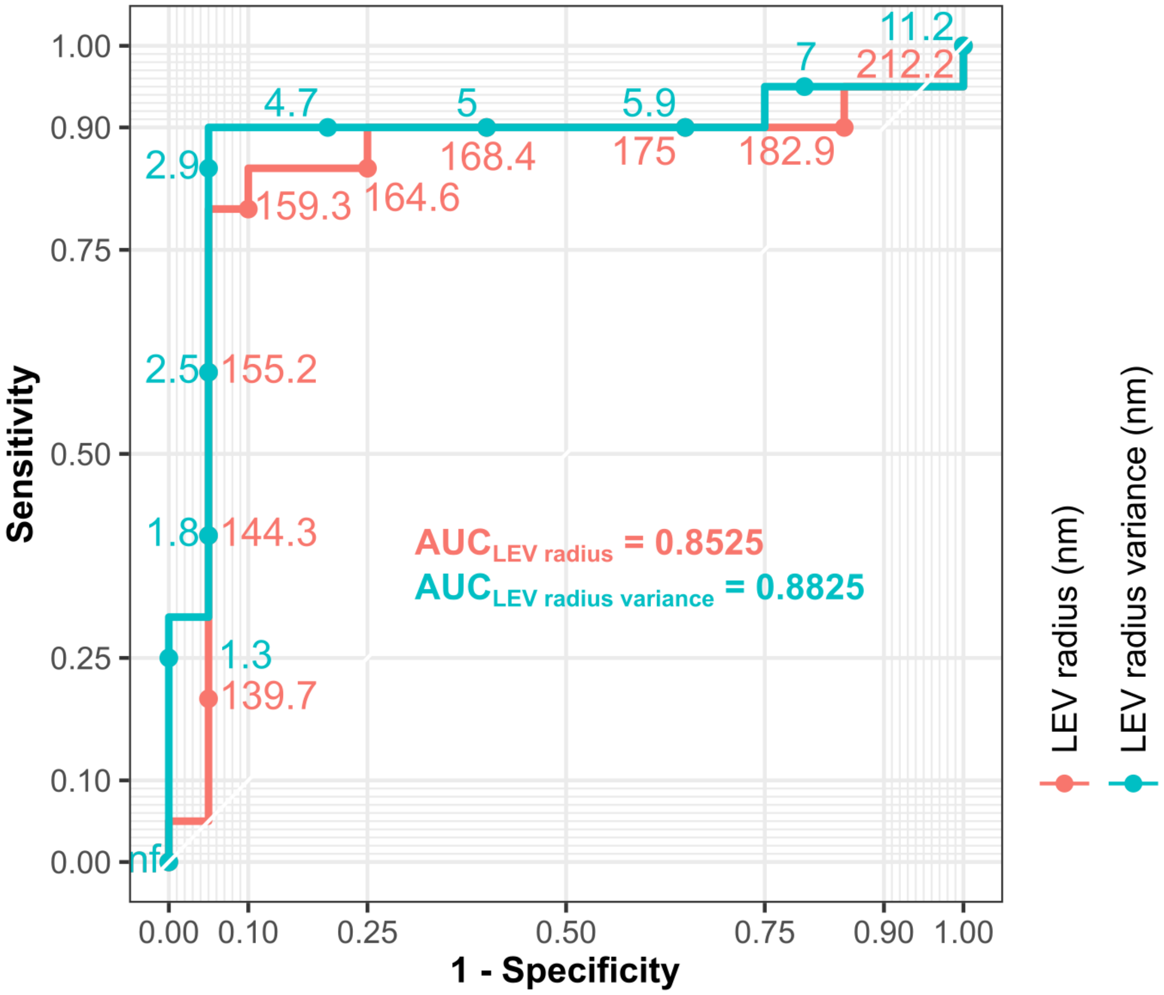

Figure S1: LEV (large extracellular vesicle) sizes and their variance as predictor for MDD (major depressive disorder)

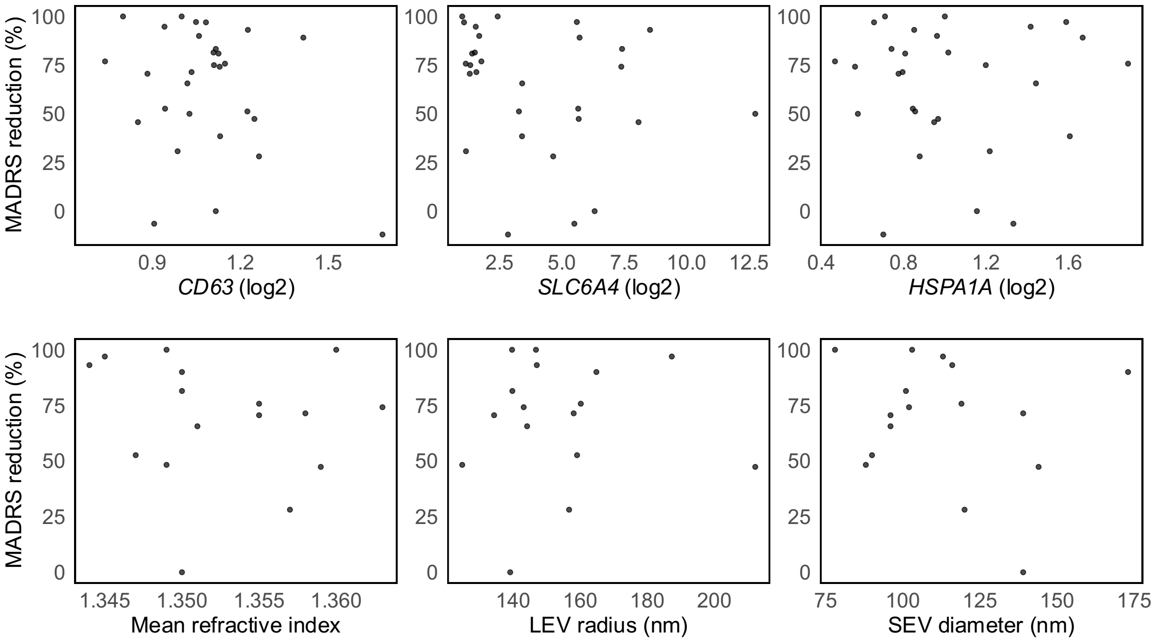

Figure S2: Scatterplots of mRNA expression and EV features in relation to antidepressant treatment response measured as the relative reduction in MADRS from screening to last follow-up visit.

#### Consort Flow Diagram

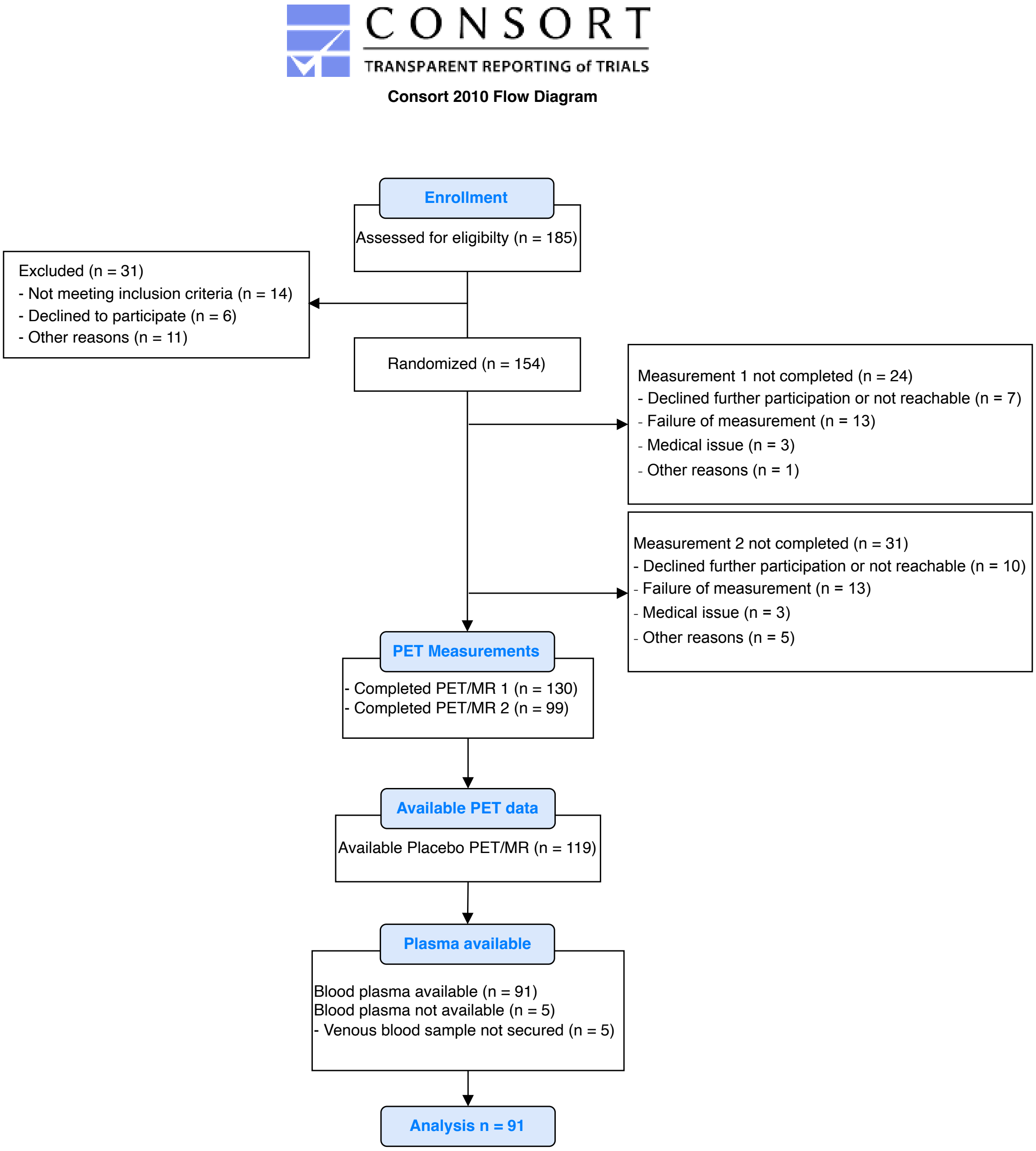
